# The heartbeat evoked potential and the prediction of functional seizure semiology

**DOI:** 10.1101/2025.07.28.25332134

**Authors:** Rohan Kandasamy, Samia Elkommos, Ineke van Rossum, David Martin-Lopez, Akihiro Koreki, Fiona Farrell, Suzanne O’Sullivan, Beate Diehl, Fahmida Chowdhury, Hugo Critchley, Matthew Walker, Sarah Garfinkel, Mahinda Yogarajah

## Abstract

Functional seizures (FS) are common, and distinguishing FS from epileptic seizures (ES) can be challenging despite video telemetry (VT), and the pathophysiology is not well-understood. The heartbeat evoked potential (HEP) reflects the central processing of cardiac signals and bodily attention. Our group previously demonstrated that HEP amplitude may differentiate FS from ES. Here, we sought to replicate these HEP findings in an independent sample and test if HEP can characterise semiological subtypes of FS and ES. Lastly, we examined whether HEP modulation was associated with real-time bodily symptom reporting in functional and vasovagal syncope.

In a first study, we identified FS or ES from VT recordings of patients. We categorised FS and ES into “motile” or “non-motile” according to semiology with predominantly positive motor features, or with subjective sensory or negative motor features, respectively. HEP amplitude was calculated by averaging EEG segments time-locked to ECG R-waves, correcting for pre-R wave baseline, to quantify the average voltage between 0.455 and 0.595 seconds after the R wave. We compared HEP amplitude across FS and ES of equivalent semiology. For the second study, we quantified the HEP amplitude in patients with functional syncope and vasovagal syncope, from EEG recorded during head-up tilt procedure, focusing on reported symptom onset rather than the onset of clinical events.

Sixty-three and sixty individuals were included in study one and two, respectively. HEP amplitude distinguished FS from ES with matched semiology: In non-motile FS, HEP become more positive at the scalp from the interictal to preictal period, whereas in motile FS, the HEP became less positive at the scalp. ES were not associated with significant changes in HEP. In individuals with functional syncope, a more positive HEP was associated with reported bodily symptoms, but not for non-bodily (psychological or emotional) symptoms. In individuals with vasovagal syncope, a less positive HEP was associated with (most) bodily symptoms.

These findings indicate that FS semiology relates to patterns of bodily attention, as reflected by HEP amplitude change. Non-motile FS were associated with preictal increased HEP amplitude, suggesting greater bodily awareness, whereas motile FS showed a decrease in HEP amplitude. The increased HEP amplitude associated with bodily symptom reporting in functional syncope further supports a role for the HEP in tracking interoceptive processing and bodily attention. Together, this highlights the potential utility of the HEP as a marker for distinguishing FS from ES and for probing the interoceptive mechanisms underlying functional symptoms.

## Introduction

Functional seizures (FS) outwardly resemble epileptic seizures (ES) but are not associated with hyper-synchronised, epileptiform brain discharges on evident on electroencephalogram (EEG). Individuals with functional seizures make up 20 to 30% of persons presenting to an epilepsy service^1^, and are associated with elevated morbidity, premature mortality^2^, and increased health economic costs^3^, comparable to those with ES alone. Despite this, and a growing body of research findings on the psycho-social co-morbidities of FS, the neurobiology of FS remains poorly understood. Correspondingly, explanatory accounts for observed inter- and intra-individual variability in FS semiology are limited. Historically, FS are subclassified along several dimensions, the predominant dimension being movement^4^: One third of FS are associated with a paucity of movement (here referred to as ‘non-motile’) whereas two-thirds of FS are associated with excessive movements (which we will refer to in this paper as ‘motile’)^5^. Functional syncope shares striking phenomenological and mechanistic similarities with non-motile FS: Both may involve transient episodes of lack of movement and/or apparent loss of consciousness without evidence of epileptic activity or cardiovascular compromise. The clinical distinction between them often reflects differences in referral pathways rather than dissociable underlying pathophysiology. Patients presenting to epilepsy services may be diagnosed with non-motile FS, whereas those seen in syncope or cardiology clinics with the same core features are often labelled as functional syncope. Functional syncope can thus be viewed as a clinically equivalent presentation to FS, shaped by service structures rather than fundamental differences in symptom generation^6^.

One of the more prominent psychological models of FS propose that there is an interplay between physiological hyperarousal, involuntary conditioned responses, and associated neural representations, that trigger the activation of a latent ‘seizure scaffold’^7^. This scaffold is structured by previous experiences and perceptions to provide a pre-conscious mental template of epileptic seizure behaviour or semiology. While this proposal has broad explanatory power, its very breadth makes it challenging to substantiate with concrete evidence. Indeed, when healthy volunteers were asked to simulate an ES, only 7% simulated a non-motile seizure, and that while there were similarities, there were also important differences between these simulated seizures and typical characteristics of FS^8^. One conclusion was that the peri-ictal experiential symptoms of individuals with FS must also therefore influence seizure semiology. The typical conceptualisation and depiction of ES in popular culture, primarily to generalised tonic-clonic (motile) seizures, is likely to inform a seizure scaffold^9^. The concept of a seizure scaffold fails to explain why some FS patients manifest individual seizures with changing semiological characteristics, for example starting with motile activity before becoming non-motile or vice versa^4^.

Models of FS based on cognitive neuroscience may better meet these challenges. Interoception is the moment-by-moment mapping of the body’s internal state at conscious and preconscious levels^10^. It is a fundamental process which is critical to the homeostatic regulation and maintenance of the bodily physiological systems that are essential for survival. We previously demonstrated using both explicit^11^ and implicit^12^ behavioural paradigms, and quantification of changes in pre-ictal EEG^13^ that both trait and pre-ictal state differences in interoceptive body-to-brain mapping are present in individuals with functional seizures. Moreover, we interpret these findings within a predictive processing framework, wherein the brain generates predictions based on past experiences (‘priors’) and then compares these predictions to actual sensory input. Discrepancies between predictions and sensory input generate ‘prediction errors’. Prediction errors are minimised by either by updating the brain’s model of the world or body (perception) or by initiating actions and behaviours to change the sensory data to fit the prediction (‘active inference’). Prediction error minimisation is further determined by the relative weighting or gain (precision) afforded to each (prediction, sensory input and error) variable. These models extend beyond proprioceptive and exteroceptive–information to interoception; Arguably, afferent interoceptive signals have primacy relative to exteroceptive or proprioceptive signals because of their fundamental role in the homeostatic regulation of bodily physiology critical to survival^14^. However, to minimise interoceptive prediction errors and maintain physiological integrity in a coordinated manner, autonomic reflexes (‘intero-action’) are often insufficient, and instead the individual is motivated to *act* upon or *move* within the world^15^. Within the framework of interoceptive predictive coding is also proposed within consciousness science to understanding and integrated ‘sense of self’: The continuous, dynamic, predictive and coordinated control of internal physiological state is the plausible substrate for a pre-conscious, minimal sense of self and bodily self-consciousness, arising from the integration of prior expectations and afferent, interoceptive sensory data^16,17^. The breakdown in this process may lead to apparent dissociative experiences. In this model, the weighting (precision) afforded to bodily signals is equivalent to the degree of (pre)conscious attentional focus upon them.

We previously reported that the amplitude of the heartbeat evoked potential (HEP) becomes less positive prior to (predominantly motile) FS^13^. The HEP is an electrocortical potential attributed to the brain’s processing of cardiac, interoceptive signals. Thus HEP provides a potential objective measure of the integrity of a core neural substrate for (pre)conscious, minimal selfhood, and its disruption in FS^18^. The HEP is modulated by attention towards or away from the body, with a less, or more positive amplitude at the scalp compared to baseline corresponding to reduced or increased attention to the body respectively^19^. We interpreted our previous findings of HEP differences in FS to a reduction in the precision weighting afforded to cardiac interoceptive prediction error signals, associated with attenuated or inhibited attention to the body linked to symptoms of dissociation and de-personalisation. We proposed that that elevated sympathetic cardiorespiratory arousal and violent movement inherent in a convulsive FS represent brain-initiated compensatory autonomic and motoric responses to this reduced precision or weighting afforded to pre-ictal, cardiac, interoceptive prediction errors. That is, a less positive HEP before a motile FS represents changes in bodily awareness, and the subsequent FS could be conceptualised as an allostatic response to the disembodied state given the fundamental assumption or ‘prior’ held with high weighting or precision (‘hyperprior’) that the brain is embodied, and there must be a body present. Here, allostasis refers to the process of maintaining homeostasis through adaptive change of the organism’s internal environment to meet anticipated or predicted demands (e.g., anticipatory rises in heart rate before exercise). Thus, within this predictive coding model of interoception, autonomic and motor reflexes which comprise a motile FS are a means of implementing allostatic control to disembodiment.

Our findings are now replicated^20^, yet they need extending beyond the semiology of motile FS. Given that it is widely accepted that excessive attention to the body or internal attention, rather than externally focused attention, impairs motor fluency in functional neurological disorders^21^, we might expect converse dynamics in attentional changes between motile and non-motile FS. Increased bodily attention is also recognised to increase or make the HEP amplitude more positive^19^. This hypothesis would also be consistent with the conceptualisation of non-motile functional seizures being a form of a freeze response^22^.

In the first study, we therefore hypothesised that 1) Motile FS will be preceded by a decrease in the HEP amplitude from baseline. 2) Non-motile FS will be preceded by an increase in the HEP amplitude from baseline. 3) Similar changes will not be seen in semiologically similar ES. In the second study. We hypothesised that spontaneous reports of bodily symptoms will correlate with increases of HEP amplitude from baseline if the increase in the HEP amplitude reflects increased bodily attention.

### Study 1: Heartbeat Evoked Potential and Heart Rate Variability in Functional Seizures

#### Materials and methods

##### Ethics

Both studies were retrospective, data-only studies (including routinely collected clinical data only). The first was conducted using NHS patient records under ethical approval granted by the NHS Research Ethics Committee (IRAS References 305776 and 331794), in conjunction with the University College London Hospitals/University college London (UCL/UCLH) Joint Research Office (Sponsors). All patient records were anonymized prior to analysis. The study complied with all relevant confidentiality and data governance guidelines, adhering to NHS policies and the regulations agreed by the UCL/UCLH Joint Research Office.

##### Patients

Patients admitted for video telemetry were identified retrospectively by final diagnosis, and then grouped by the seizures captured on telemetry (FS, ES). All final diagnoses were based on the review of the telemetry and other clinical data and the consensus opinion of both the reporting neurophysiologist and neurologist in charge of the patient’s care. We did not include patients with equivocal diagnoses, nor did we include patients who did not have events during the recording (or who only had events before EEG leads were applied). Events were reviewed both in the consultant report and by visual inspection of the video record to identify and categorise the events semiologically. ES were initially categorized according to the ILAE semiological classification.

To permit clear comparison, the events (both FS and ES) were grouped into broad categories: ‘motile’ events, which, referring to both FS and ES that had predominantly positive motor features; ‘non-motile’ events, which referred to FS and ES with either predominantly negative motor features or subjective symptoms and minimal/no movement. This allowed comparison of events between FS and ES that had broadly similar semiology to assess the utility of the HEP as a diagnostic tool. Focal motor and generalized motor FS were classified as motile, while akinetic and those patients presenting primarily with reported subjective symptoms with minimal or no overt movement (eg. altered awareness, internal sensations, or speech arrest) were classified as non-motile. FS with semiologies that varied in semiology or which were not clearly categorizable, were excluded. Examples of ES semiological classifications classified as motile include: generalised tonic-clonic, hypermotor, focal motor with/without preserved awareness, automotor. Examples of non-motile ES include: dialeptic, absence, as well as isolated auras.

##### ECG and EEG recording

EEGs were recorded using standard 10-20 montages. Some recordings had supplementary electrodes, but these electrodes were not included in the analysis. The minimum electrodes for inclusion were Fz, Fp1, Fp2, F3, F4, F7, F8, C3, Cz and C4. Contemporaneous ECG recording was also required for inclusion. Only 1 ECG lead was included in the analysis, though at times more were available. If several leads were recorded, the clearest (i.e. least affected by artefact) lead was recorded.

EEG signals with a sampling frequency of 256Hz were used, with digital maxima and minima of +/- 32767. High and low pass filters of 0.53 and 70Hz, respectively, were applied digitally. Suitability for inclusion was assessed with the EEG in a longitudinal bipolar montage and an average montage prior to extraction of the EEG data. The recordings were exported as anonymized EDF+ files.

##### Events

Events were identified from the report and from annotations in the EEG record, and then examined in the video record. They were excluded if less than five seconds long, or if there was no concurrent EEG and ECG recording (for example for events that occurred before lead application). They were excluded if the event was equivocal (i.e. the reporting consultant did not conclude the event was either ES or FS).

##### EEG Segment Selection

Three types of EEG segments were selected: interictal, preictal, and postictal. Interictal periods were manually selected as periods of relatively artefact-free segments of wakeful background activity – with a minimum duration of five minutes. The onset and offset of events were identified. In the case of seizures with EEG changes, the onset was set at the point of the onset of either the EEG or the clinical seizure depending on what occurred earlier. In the case of non-motile seizures, if there was no EEG change, behavioural arrest, or report of onset by patient (whichever is earlier) was used as the onset time. In motile events without EEG change, onset was marked by the onset of motor activity or cessation of normal behaviour, whichever was earlier. As with our previous methods^13^, up to five minutes preictally and postictally was selected for each event. Seizures were grouped by patient, by semiology (motile or non-motile), and by type (epileptic or functional). When a patient had more than one seizure of the same semiology and type, the data were averaged to produce a single value for that combination. Otherwise, the methodology for selecting clips reflected the methods used in the previous paper.

##### Artefact Correction

To compensate for artefacts that may have obfuscated or artificially increased the HEP several techniques are available. In the present study, artefact subspace reconstruction was used^23^ with a threshold of 20.

To reduce the risk that the subsequent R wave would cause an electrocardiographic artefact that would increase the HEP, we removed R-R intervals from averaging that were too brief (<700ms). This reduced the number of R-R epochs included in the average, and if too few remained to reliable derive a reliable average (less than 60) the analysis automatically excluded these EEG segment averages from analysis.

##### HEP derivation

The HEP is derived from measuring the voltage in a segment of EEG segments back averaged according to the R waves of the ECG. We averaged segments from 0.4 seconds before the R wave to 0.8 seconds after the R wave. Epochs wherein the maximum potential difference exceeded 100µV were rejected from the averaging, to prevent large amplitude graph elements and artefacts from distorting the average^24^. Once these segments are back averaged, the baseline is corrected to the baseline before the R wave – in this experiment we used between 0.35 and 0.1 seconds prior to the R wave as the baseline. The average voltage of the segment of the averaged epochs between 0.455 and 0.595 seconds after the R wave was calculated, in accordance with previous work^25^. This average potential difference was computed for all leads included in the analysis. To reduce the number of statistical inferences (and hence reduce the multiple comparisons problem), we limited our analysis to the ‘compound HEP’, the sum of the leads identified as the most significant in previous work (C4 and F8)^13^.

##### Heart Rate Variability

Heart Rate Variability (HRV) was computed using the neurokit2 module^26^, using the raw ECG leads as input. We chose the root-mean-squared successive difference (RMSSD) as the measure of choice HRV (RMSSD) is recognized as an index of cardiac parasympathetic drive^27–29^ linked to baroreflex function. Heart rate variability (HRV) was computed from the raw ECG using the neurokit2 module^26^, using the raw ECG leads as input. We selected the root-mean-squared successive difference (RMSSD) as our primary HRV measure, because it is a well-established index of cardiac parasympathetic drive^27–29^ closely linked to baroreflex function. RMSSD is sensitive both to **bottom-up interoceptive signals from baroreceptors,** which detect changes in blood pressure, and **to top-down influences on baroreflex control** exerted by the brain. A reduction in RMSSD thus reflects diminished vagal modulation, allowing heart rate and blood pressure to rise together, a pattern characteristic of states of cardiovascular arousal. No definite measures of sympathetic activation are available from HRV alone^30^; low- and high-frequency powers, show some modulation by sympathetic activity, but are also influenced by parasympathetic activity. With this caveat, we included these, and heartrate as proxy measures of sympathetic activation.

##### Statistical Analysis

Sample sizes were based on our group’s previous work^13^, doubled to permit division by semiology. Statistical analysis was preliminarily performed using JASP^31^, except where bootstrapping was necessary, in which the statsmodels^32^ and scikit-learn^33^ python modules were used. The normality of data was assessed visually with QQ-plots. Outliers were automatically excluded if they were more than three standard deviations away from the mean.

Comparing the effect of semiology on the change in HEP from inter- to preictal, and to take into account the heteroskedastic distribution of events between individuals, a Linear Mixed Model was not applicable due to singular fit occurring with random effect variables. Instead, to account for within-subject dependencies and unbalanced group structure, we applied non-parametric resampling at the subject level, stratified by seizure type and semiology where applicable. Inference was based on bootstrapped F-statistics, from which empirical p-values were derived. Using this method, two ordinary least square analyses were performed to compare the effect of the independent variables of interest over and above the effect of the null variable (change in heart rate). Equivalently bootstrapped two-tailed unpaired t-tests were used, with empirical p-values. For within-individual testing of the significance of changes in the HEP, bootstrapped one-sample two-tailed t-tests were used.

## Results

### Demographics

Demographics are summarized in Table 1.

**Table 1:** Demographic details. Note: Some patients had seizures of different types or semiologies.

| Seizure Type | Semiology | Number of Patients | Mean Age (SD) |
| --- | --- | --- | --- |
| Functional Seizures | Motile | 30 | 35.93 (11.22) |
|  | Non-Motile | 27 | 39.76 (15.25) |
| Epileptic Seizures | Motile | 22 | 42.05 (17.26) |
|  | Non-Motile | 9 | 41.22 (19.33) |
| Total Number of patients |  | 63 | 38.0 (14.48) |

### Changes in HEP over time, in ES and FS

Performing an ANOVA across all data from both seizure types, there was a significant interaction of epoch (i.e. interictal, preictal and postictal periods), seizure type and semiology (F 8.396, p<.001) as well as a significant but smaller effect of Epoch * Semiology (F=3.832, p=0.024) and Epoch alone (F=3.632, 0.029). There was a significant interaction of epochs and semiology in FS (F=10.0, p < 0.001). There was no effect of this interaction on ES (F=2.030, p=0.141). These changes over time are shown in Figure 1. In this unpaired analysis (i.e. successive HEP values were not grouped within individuals) post-hoc tests were insignificant after alpha adjustment.

**Figure 1:**
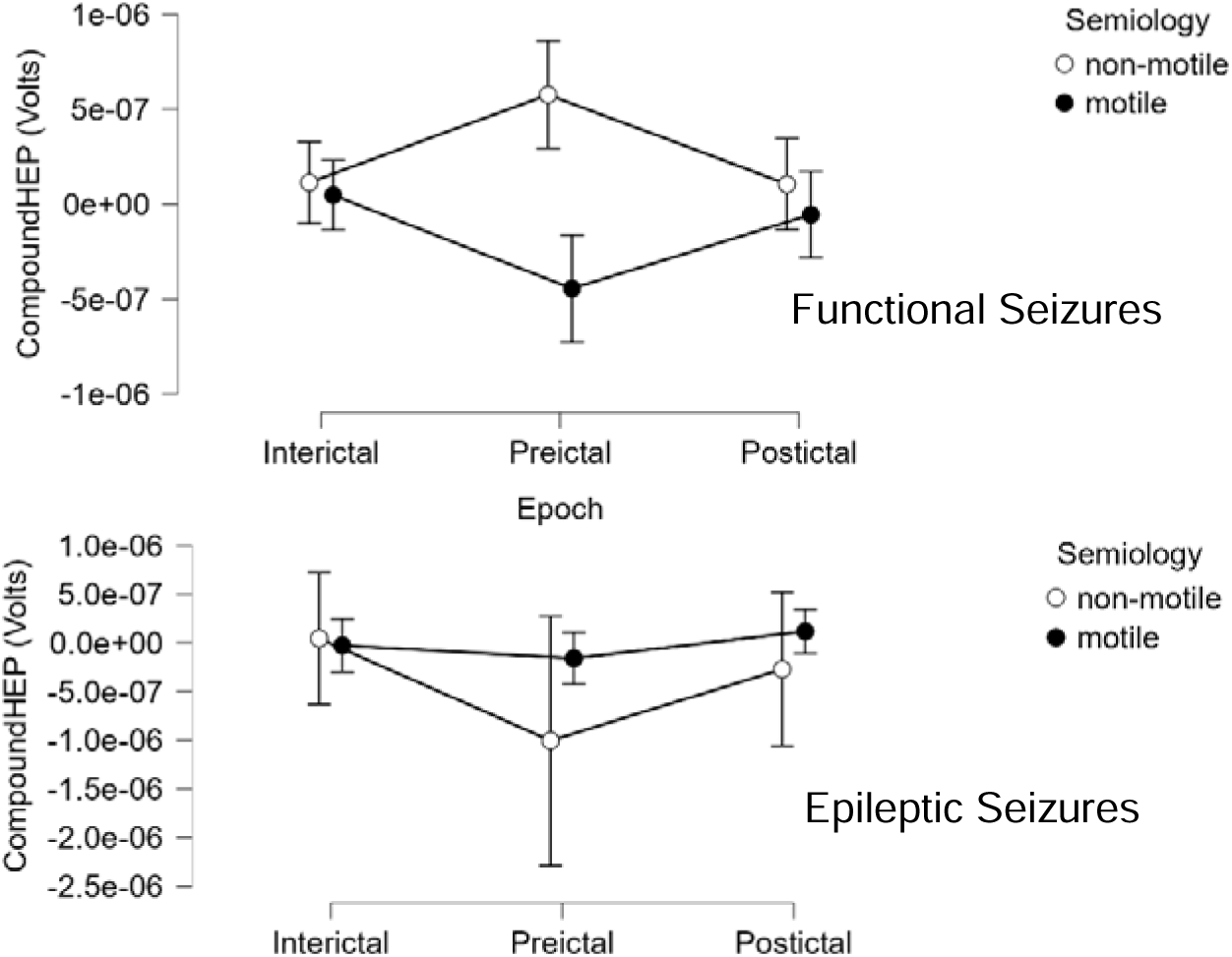
Change over time for Compound HEP. (the sum of Heartbeat-Evoked Potentials at C4 and F8 leads) over different epochs around functional seizures (top) and epileptic seizures (bottom).

In order to demonstrate significant changes in the compound HEP in a given individual with seizures of different types or semiologies, we computed the difference between the HEP in the interictal and preictal period within an individual (‘Compound HEP Change’). There was a significant effect of semiology on these changes in the FS group, over and above the effect of any changes in heart rate (F = 14.9, p < 0.001, Bootstrapped (mean) F = 16.2, empirical p-value (EPv) < 0.001). There was no effect on the Epileptic Seizure group (F = 2.80, p = 0.11).

There was a significant Compound HEP decrease in motile FS (T=-3.6, p=0.0012, EPv < 0.001, alpha=0.0125), and a significant increase in non-motile FS (T=4.0, p=0.002, EPv < 0.001). There were no significant changes in the ES of either semiology (motile: T = - 0.050, p = 0.9605; non-motile: T = -2.25, p = 0.031). These effects are summarized in Figure 2.

**Figure 2:**
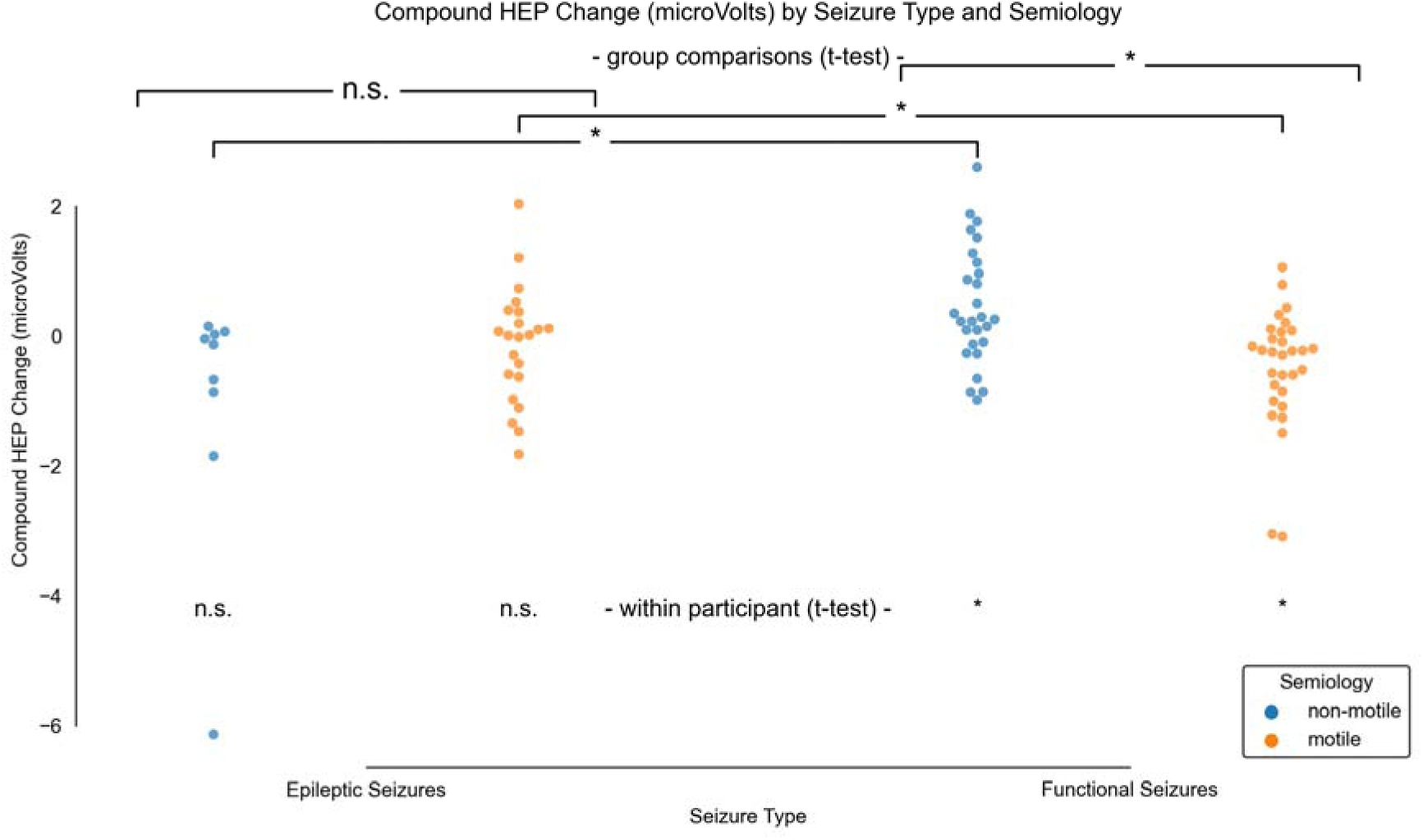
Changes from the interictal to preictal periods for both seizure types. (functional and epileptic) for different semiological groups (motile and non-motile). “*” marks a significant finding in the bootstrapped t-tests; n.s. = non-significant. “Within participant comparison” refers to the paired t-test comparison of the interictal and preictal period for a given participant and event type

### Between-group analysis of HEP change – motile ES v motile FS & non-motile ES v non-motile FS

Figure 2 shows the group-wise comparison of the change in Compound HEP. Comparing seizures of matched semiological groups, but different aetiologies (ES vs FS), there was a significant difference between motile FS and motile ES after Bonferroni correction (T=-2.6, p=0.011, EPv < 0.001, alpha=0.0125), and between non-motile FS and non-motile ES (T=2.8, p=0.0074, EPv < 0.001). Comparing seizures of the same type (i.e. ES or FS) but with different semiologies, there was a significant difference in the HEP change when comparing motile and non-motile FS seizures (T=4.50, p<0.001, EPv = <0.001). There was no significant difference in the change in compound HEP values when comparing motile and non-motile ES (T=1.7, p=0.10, EPv = 0.054).

### Heart Rate Variability changes within FS groups – motile v non-motile

At the interictal baseline, patients with motile FS had higher RMSSD (T=4.1892, p-value< 0.001, EPv=<0.001) than those with non-motile functional seizures. This difference was no longer significant pre-ictally. The difference in the change in interictal RMSSD to preictal RMSSD between motile and non-motile functional seizure groups was not significant, though there was a trend for a greater increase in RMSSD in non-motile FS (T=2.0, p=0.05 - uncorrected). There were no differences in interictal, or the change in interictal to preictal LF or HF power between motile and non-motile FS.

### Excluding Cardiodynamic Confounds

The ANOVA comparing the interaction of seizure type and semiology was robust to comparison with a null model that incorporated changes in the heart rate. In the FS group, the motile group had a negative change in the heart rate preictally, and the non-motile group had a positive change preictally, though this was not significant (p_holm_ = 0.257 and 1, respectively, see Figure 3). The Interictal heart rate was higher in the motile group (T = 8.0032, p-value < 0.001, EPv < 0.001). Correlation analysis of the change in HEP and the change in heart rate did not show a significant correlation (Pearson r = 0.125, p = 0.25), lending support to the conclusion that these changes in HEP are not due to changes in heart rate. The absence of significant correlation may partly reflect the heart rate thresholding method we used to prevent the next R wave from introducing field artefact into HEP computation.

**Figure 3:**
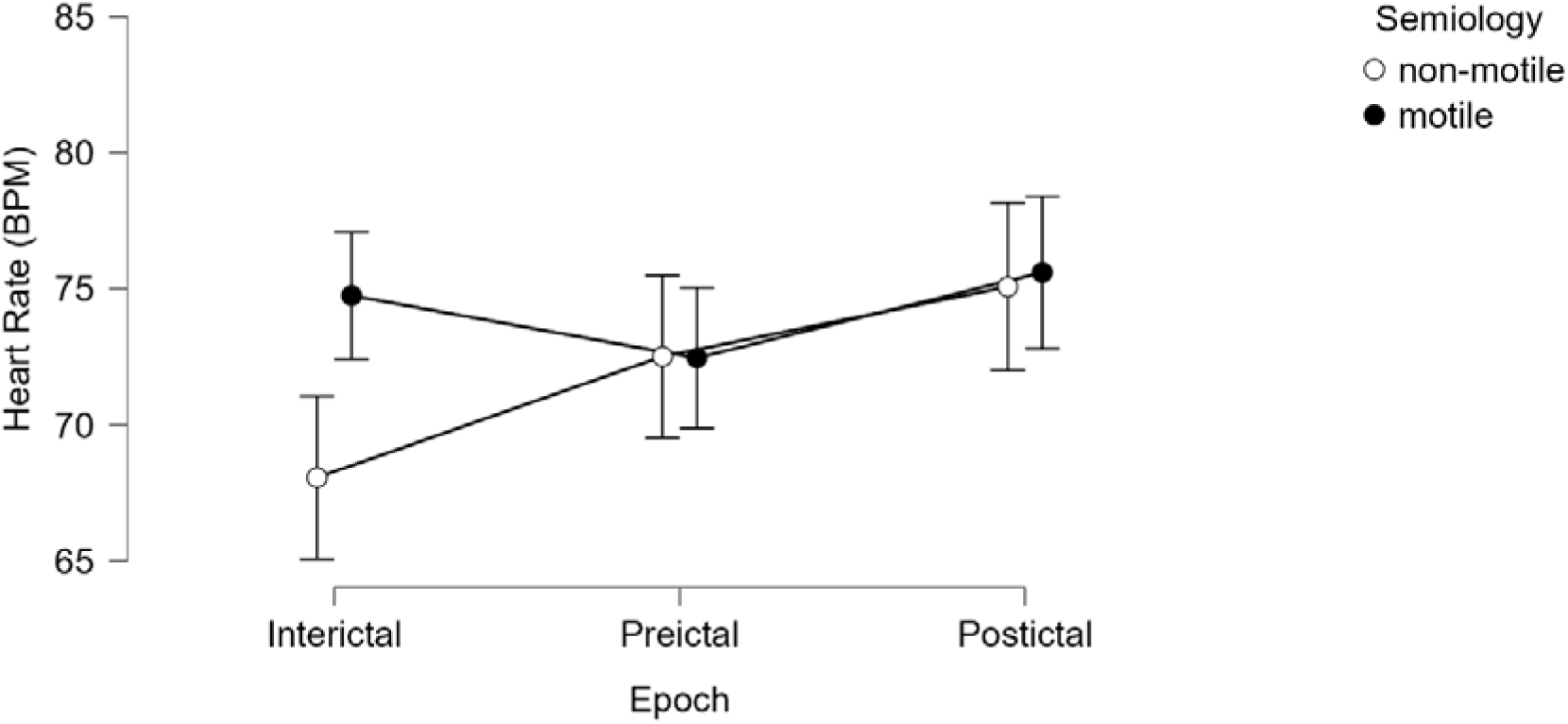
Heart rate at different periods (functional seizures).

### Experiment 2: Symptom-Linked HEP in Functional Syncope: A Naturalistic, Independent Cohort Reflecting Non-Motile FS

The goal of this second experiment was to investigate the changes in HEP associated with symptom reporting, with the hypothesis that an increase in somatic symptoms would be associated with bodily attention, and hence an increase in the HEP.

#### Methods and Materials

##### Ethics

For the second study (based at Leiden University Medical Centre (LUMC), Leiden, the Netherlands), the use of anonymized data was approved by the appropriate institutional governance procedures, in accordance with local regulations and ethical standards and Dutch law.

###### Data selection and processing

Simultaneous EEG and ECG recordings were taken from age and sex matched sets of 30 patients with functional syncope and 30 with vasovagal syncope occurring on tilt-table testing. The benefit of the tilt-table investigation over and above typical formats for video telemetry is the attendance of a clinician throughout the test, which permits contemporaneous reporting and documentation of symptoms reported by the patient prior to the functional syncope, which are recorded as annotations in the data.

The methods duplicated those of the previous experiment, but with the caveats that no interictal EEG/ECG data was available due to the nature of the test, and that due to high heart rates associated with the procedure, very few epochs were suitable for inclusion. To allow comparison between periods wherein symptoms were reported versus not reported, the data was stratified by patient, and by whether the epoch was in the two minutes preceding a symptom occurring. Symptoms were categorized into cardiac (e.g., palpitations), abdominal (e.g., nausea), respiratory (e.g., shortness of breath), cephalic (e.g., lightheadedness or vertiginous symptoms), peripheral (e.g., tingling extremities) or psychological/emotional symptoms (e.g. fear). From these stratified datasets bootstrapped sampling was performed to allow inclusion of sufficient epochs to permit meaningful averaging – with 20 bootstrapped averages for each patient. Alpha was set at 0.0083 to account for multiple comparisons (6 symptom groups). Normal Student’s t-tests were used to compare between the epoch groups.

### Results

In the patients with functional syncope, there was an increase in the HEP at times where the patient was reporting cardiac, cephalic, abdominal, respiratory or peripheral symptoms, from baseline (Figure 4). There was no significant change when reporting psychological or emotional symptoms (such as panic or anxiety), suggesting that this is a specific phenomenon to ‘bodily’ symptoms, and is also not an artefact of speech. Interestingly, during cardiac, cephalic, respiratory or peripheral symptoms, there was a significant decrease in the HEP in the patients with vasovagal syncope (Table 2). This divergence suggests that HEP dynamics are not reducible to symptom type or speech artefacts, but reflect fundamentally different underlying mechanisms. In functional states, HEP modulation likely reflects attentional or representational processes, whereas in vasovagal syncope, it may reflect afferent signal suppression or reduced cortical responsiveness due to autonomic collapse.

**Figure 4:**
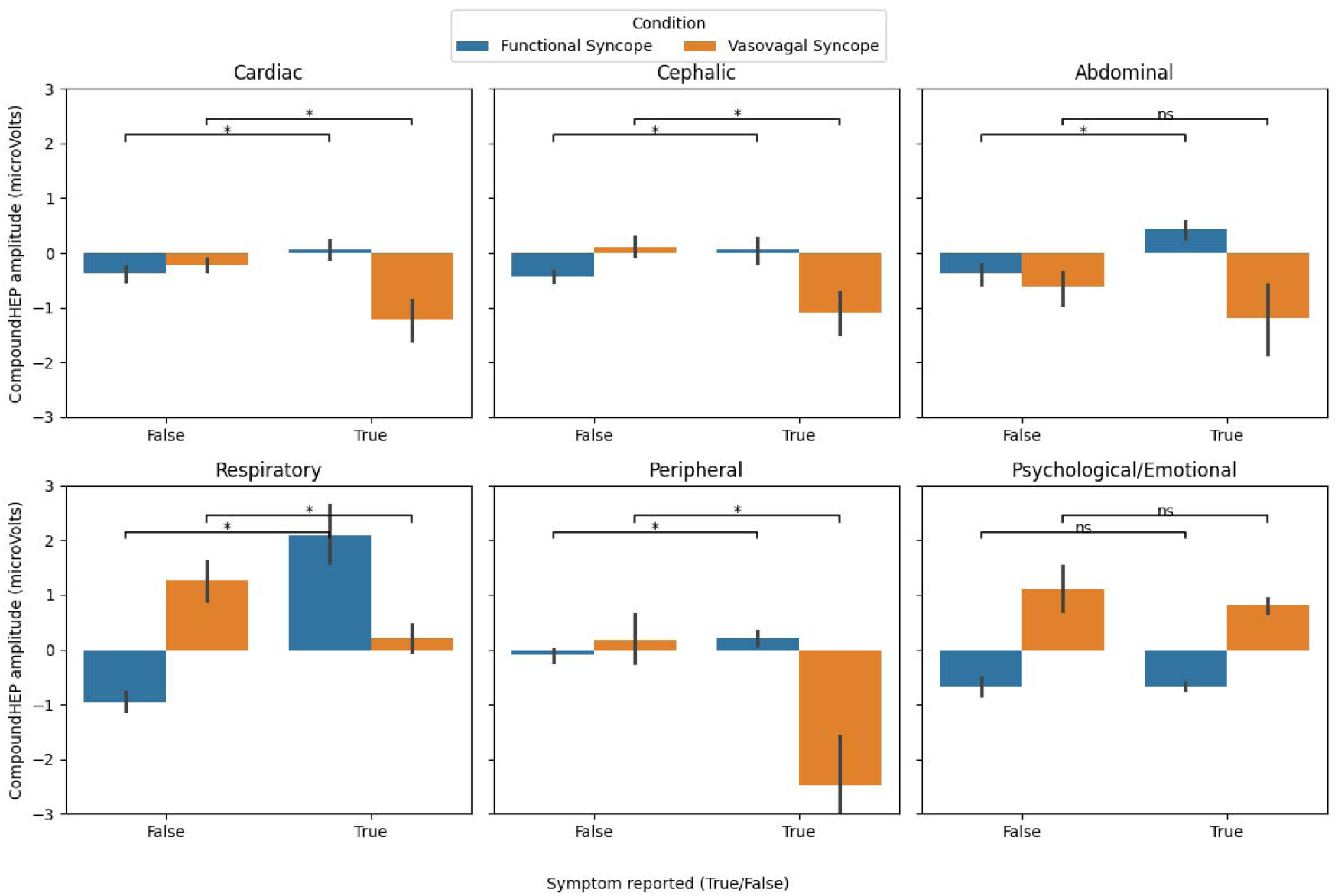
Compound HEP (Heartbeat evoked potential at C4 and F8 summed), compared between epochs within 120 seconds of a symptom being reported, versus those without. * = significant, n.s. = non-significant.

**Table 2:**
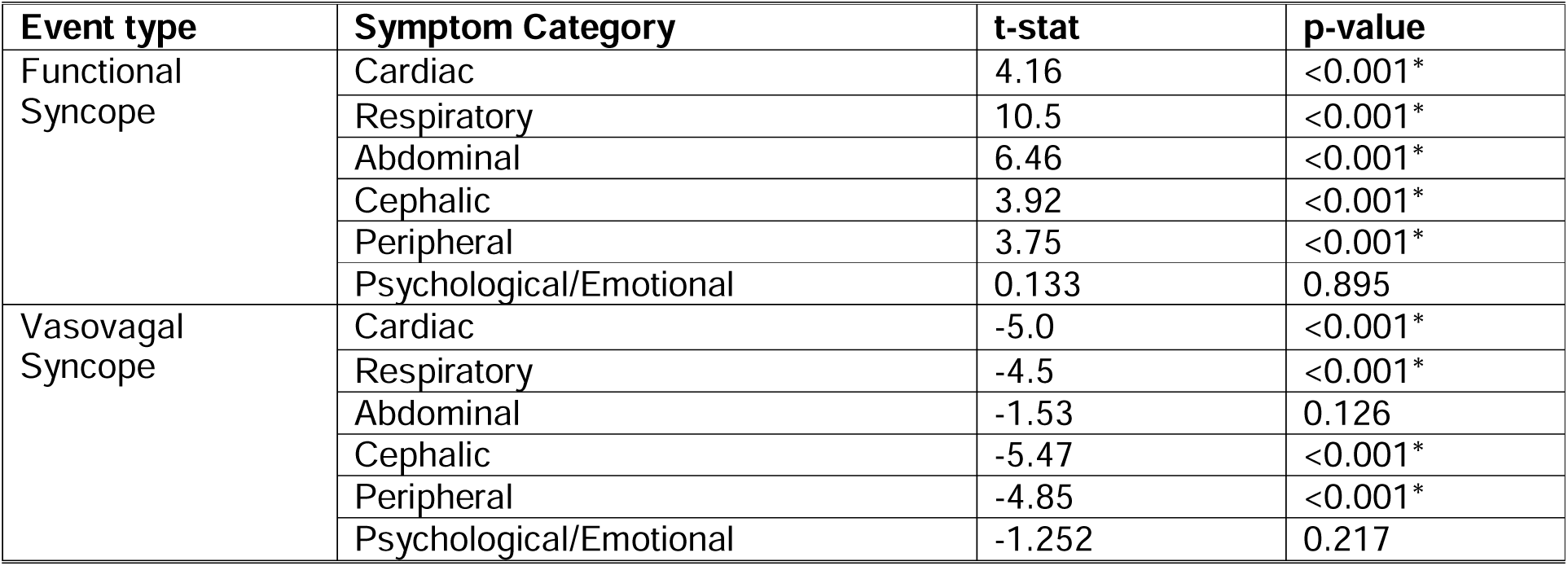
Statistical comparison of heartbeat evoked potential in segments proximal- or non-proximal to symptom reporting. *=significant.

| **Event type** | **Symptom Category** | **t-stat** | **p-value** |
| --- | --- | --- | --- |
| Functional<br>Syncope | Cardiac | 4.16 | <0.001* |
|  | Respiratory | 10.5 | <0.001* |
|  | Abdominal | 6.46 | <0.001* |
|  | Cephalic | 3.92 | <0.001* |
|  | Peripheral | 3.75 | <0.001* |
|  | Psychological/Emotional | 0.133 | 0.895 |
| Vasovagal<br>Syncope | Cardiac | -5.0 | <0.001* |
|  | Respiratory | -4.5 | <0.001* |
|  | Abdominal | -1.53 | 0.126 |
|  | Cephalic | -5.47 | <0.001* |
|  | Peripheral | -4.85 | <0.001* |
|  | Psychological/Emotional | -1.252 | 0.217 |

## Discussion

We demonstrate that in individuals with motile and non-motile FS, HEP amplitude becomes significantly more negative and positive respectively before the seizure. To our knowledge, this is the first study ever to demonstrate objective neurophysiological differences between semiological subgroups of FS. Given the retrospective and clinical nature of this study, we are limited in the direct inferences that can be drawn about what these differences reflect symptomatically. However, given that in individuals with motile and non-motile ES, the HEP does not significantly change before the seizure, we infer that these changes are not directly related to the presence or absence of motor activity during the seizure. Instead, we propose that differences in attentional direction *prior* to a seizure give rise to the differences in the HEP amplitude. Thus, we propose that seizure semiology (motile vs non-motile) reflects whether the brain is attempting to resolve interoceptive uncertainty through action (motile FS) or through suppression and shutdown (non-motile FS), mediated by the attentional weighting of bodily signals before the event.

### HEP as an index of bodily attention and interoceptive precision

A recent meta-analysis of HEP studies identified several factors that modulate the HEP amplitude^19^. These include arousal, the underlying clinical condition in an individual and the direction of an individual’s attention at the time of testing. In this study, we have the advantage of studying a group of participants who share the same clinically defined condition of functional seizures for whom repeated within-participant HEP measurements were recorded at different time points, that is, both between and before their seizure. In our analysis, we applied strict R-R interval thresholding to ensure minimal cardiac field contamination of the HEP, and showed that in this ‘thresholded’ data there were no arousal differences between interictal and pre-ictal periods across groups, as indexed by heart rate. Finally, linear regression analyses of the change in HEP amplitude included heart rate as a co-variate of no interest. For these reasons, we are confident the observed and reported differences here are not due to changes in arousal or differences in the underlying clinical condition. Instead, we propose that the differences in HEP amplitude reflect differences in the directionality of attention before the seizure, namely increased inwardly towards the body or directed away from the body.

One of the earliest studies to test whether attentional focus modulates HEP amplitude used a paradigm contrasting interoceptive and exteroceptive attention and found that directing attention towards the body increased HEP amplitude, making it more positive^34^. This supported the view of the HEP as a neural correlate of interoceptive prediction error associated with each heartbeat^35^. Interoceptive or inwardly directed attention to the body, enhances the relative precision of interoceptive signals, increasing the weight of associated prediction errors, while outward attention reduces their salience. The increased precision of interoceptive prediction errors results in their propagation up the predictive hierarchy to update models for more accurate future predictions about each heartbeat. This precision-modulation framework interpretation also explains frequently reported findings of a positive correlation between the HEP and cardiac interoceptive accuracy scores on the heartbeat tracking task^36,37^. Those individuals who can better direct attention to their internal bodily states will be able to increase the precision of cardiac interoceptive signals and will therefore have a greater or more positive HEP amplitude, as well as greater cardiac interoceptive accuracy scores^35^.

These findings have been extended by studies using paradigms involving, omitted tones^38^, where heartbeat-synchronous expectations about auditory stimuli result in HEP modulation when tones are unexpectedly absent. These effects were amplified by inward attention, further supporting the link between interoceptive focus, increased prediction error precision, and enhanced HEP positivity.

An alternative, but complementary interpretation is that the HEP represents a core aspect of the ‘self’, arising from the brain’s preconscious and continuous neural monitoring of intrinsic, spontaneous bodily signals such as those pertaining to the heart^39^. Studies using MEG^40^ and intracranial EEG^41^ have shown that imagining scenarios from a first-person (vs third-person) perspective increases HEP amplitude, supporting its role as a neural index of embodied subjectivity. While our study lacks structured self-report measures of attention, increases in HEP were consistently seen contemporaneously with bodily (but not emotion) symptom reporting in the second experiment, specifically in the individuals with functional syncope. This suggests that these HEP changes are unlikely to be mere artefacts of speech or arousal, and instead and instead likely reflect shifts in bodily attention within a first-person experiential frame, that is, pre-reflective awareness of internal physiological signals. This may help explain why, in study 2, emotional symptoms, which are also first-person but not purely bodily focused, did not show similar HEP increases.

### How interoceptive precision shapes motor control

If HEP modulation prior to a FS represents shifts in bodily attention and interoceptive precision, then why do seizures differ in associated movement? This may be explained by growing evidence linking interoception and motor control. The original predictive processing model of interoception proposed a close relationship between presence, interoception, movement and agency^16^. Every voluntary action involves not only motor commands but also predictions about its interoceptive consequences, such as changes in heart rate, muscle tone, or breathing. These predictions support both agency (subjective feeling of being the causal origin of one’s own actions and their consequences) and presence (the subjective feeling of being embodied). While agency depends more on accurate sensorimotor predictions and presence more on interoceptive coherence, both rely on the brain’s ability to integrate predicted and actual bodily signals across these channels.

Other models extend this to allostasis, where interoception supports homeostasis by adjusting internal states in anticipation of future needs. While this can be mediated in part via the resolution of interoceptive prediction errors via autonomic reflexes, the resolution of exteroceptive and proprioceptive prediction errors at different time scales also contribute. That is, movement is critical and only when an individual can detect their current bodily needs, can they adjust behaviour accordingly. Signals from the viscera are therefore linked to actions in various contexts including exploration, food-seeking behaviour and fight/flight responses^15^. More recent models emphasise a bidirectional loop between interoception and movement: motor actions shape interoceptive states, and interoceptive predictions regulate motor behaviour^42^. Motor actions or movement generate expectations about internal bodily consequences and states (e.g. changes in arousal) mediated by the autonomic nervous system. Conversely, ongoing interoceptive and autonomic signals continuously influence the initiation and control of movement. Reflexive movements—conscious, adaptive, and error-monitored— depend on interoceptive input, whereas pre-reflexive movements occur automatically, without the need for interoceptive input or conscious feedback or correction.

Experimental evidence supports this interaction between interoception and motor control. TMS studies show that corticospinal excitability peaks during systole, particularly when neural responses to heartbeats—or HEP amplitude—are stronger^43,44^. During systole, baroreceptors in the aortic arch and carotid sinus are activated by rising blood pressure and send afferent signals to the brain that influence ongoing cortical activity. These signals are thought to influence cognitive processing by competing for limited attentional resources. To accommodate this rhythmic input, the brain dynamically adjusts the precision weighting of interoceptive and exteroceptive signals. During systole, interoceptive signals are assigned greater precision, supporting motor readiness but reducing perceptual sensitivity. During diastole, precision shifts toward exteroceptive signals, enhancing perception^45^. These oscillations may serve adaptive functions: lowering heart rate to prioritise perception in freeze responses, or raising it to facilitate action in fight-or-flight states^45^.

### Motile seizures

While cardiac systole has a cyclical excitatory effect on baseline motor cortex activity, behavioural event related potential (ERP) studies show that interoceptive signalling is also crucial for effective motor control. In a reward-based task, participants who could pre-consciously anticipate outcome-related bodily states—reflected in higher anticipatory HEPs— showed enhanced motor preparation (increased CNV amplitude) and faster responses^46^. This aligns with findings that lower interoceptive perceptual accuracy is associated with stronger urges to move^47^ and higher trait impulsivity^48^, suggesting that impaired interoceptive feedback may permit disinhibited, pre-reflexive motor actions. It is also consistent with research showing that external attention, enhances motor performance more than internal attention, in part because of increased corticospinal and intracortical excitability^49,50^.

Thus, in motile FS, reduced HEP amplitude may reflect diminished bodily attention and interoceptive precision, allowing the highly weighted priors of embodiment to drive involuntary, unregulated movements for which individuals report no agency. This interpretation is supported by associations between reduced interoceptive perceptual accuracy^51^, or noisier interoceptive input^52^ and diminished agency. Clinically, patients often also report relief post-seizure^53^ and experimental studies show that in the classical rubber hand illusion, the sense of agency and embodiment over the rubber hand can be enhanced by movement^54^. We therefore propose that motile FS represent a maladaptive, regulatory response to disrupted interoceptive self-representation, where unregulated, pre-reflexive movement acts as a policy for re-establishing embodied presence.

### Non-motile seizures

Non-motile seizures may also reflect a form of dysregulation of interoceptive – motor coupling, but in the opposite direction. In a study using a stop signal reaction task a go cue was unpredictably followed by a stop signal requiring the cancellation of the prepotent response^55^. Interoceptive processing, specifically during cardiac systole, disrupted response inhibition. The study found prolonged stop-signal reaction times, reduced stop-signal P3 amplitudes (an event related potential reflecting neural activity associated with inhibitory control), and higher heartbeat-evoked potential amplitudes when the stop signal coincided with cardiac systole. This suggests that a finite attentional resource shifts at a pre-conscious level toward internal bodily signals during systole as indexed by the increased HEP signal, impairing external motor control and inhibitory processes. This idea has been corroborated more directly by using EEG to show that when visual events are repeatedly timed with strong internal signals (heartbeats), the brain gradually reallocates attention toward the internal domain. This was reflected in increased HEPs and decreased visual processing, as measured by steady-state and event-related visual potentials^56^. Their findings support the idea that attention at both is a limited resource, and sustained internal focus comes at the expense of external awareness—demonstrating a longer-term trade-off between interoception and exteroception. The attentional shifts occurred preconsciously, as participants were unaware of the heartbeat coupling and were not instructed to focus on internal signals, yet the neural measures revealed a clear trade-off in processing resources.

Based on this premise of a finite attentional resource balanced between internal and external attention, we propose that excessive interoceptive representation in the brain reflected in an elevated HEP and mediated by increased bodily attention leads to dysregulated interoceptive-motor coupling, and over-regulation of motor systems and a non-motile seizure presentation. Here a rapid and overwhelming increase in the precision of interoceptive prediction errors that is too large to resolve through typical means, leads to the allostatic response or policy of triggering of an inhibitory response, where the brain ‘shuts down’ motor activity to minimize further errors or protect against perceived salient, internal threats.

This response could also be conceptualised as a freeze response. A freeze response is defined as a special case of threat-induced motor inhibition whose evolutionary purpose is to avoid detection by predators and to enhance perception. In addition to immobility, it is defined by parasympathetic dominance relative to sympathetic control of the heart, although this may not be total and there may therefore not be an absolute bradycardia. This is in contrast to sympathetically dominated fight-or-flight responses^57,58^. Motile FS showed higher vagal tone at baseline (interictally), but this group difference was no longer present preictally, suggesting a state shift in non-motile FS toward parasympathetic dominance. While the interictal-preictal difference in RMSSD change did not reach statistical significance, the trend toward a greater increase in the non-motile group (T = 2.0, p = 0.05) further supports the possibility that these seizures are preceded by autonomic profiles consistent with the interpretation of non-motile FS being similar to a freeze response. This model of FS has some parallels with the polyvagal theory which frames bodily responses to threat along a spectrum of autonomic states. In polyvagal theory, the ‘freeze’ response (non-motile FS) is linked to vagal mediated shutdown, while ‘fight or flight’ (motile FS) corresponds to sympathetic arousal^59^. The few behavioural studies that have looked at freeze responses in humans also corroborate this interpretation of the data. A study of 404 non-clinical participants found that heightened attention to bodily sensations, measured via anxiety sensitivity, predicted freeze responses during a CO₂ inhalation challenge, even after controlling for trait anxiety. Around 13% reported significant immobility, despite the absence of real threat. This supports the idea that hypervigilance to bodily sensations alone can provoke a freeze response^60^. This heightened attention to the body would also have the secondary effect of leading to failure of attenuation of sensory prediction errors that is typically needed for movement^61^. Within predictive coding models movement normally only occurs when proprioceptive prediction errors at the spinal cord level generated by confident high-level sensorimotor predictions or expectations are precise, in comparison to the precision of ascending sensory prediction errors which convey evidence against the prediction that one is moving and are normally attenuated. Action therefore requires ‘dis-attention’ form the body, and pathological attention to the body is recognised to cause immobility^62^.

### Predictions of data interpretation

In summary, the data presented here suggest that there is an optimal range of interoceptive feedback that is needed for efficient and controlled motor actions. In individuals with FS this feedback may breach this range, and the precision of interoceptive prediction errors may rapidly increase or decrease due to be increases or decreases in conscious and preconscious bodily attention prior to the seizure.

This proposed mechanism would predict that individuals with greater dynamic changes in reported somatic symptoms (reflecting greater bodily attention as indexed by the change in the HEP) prior to a seizure may be more prone to non-motile seizures. Conversely, those individuals with greater dynamic changes in reported dissociative symptoms (reflecting reduced bodily attention as indexed by the change in the HEP) may be more prone to motile seizures. This prediction has been proved in part by trait studies highlighting that individuals with ‘pauci-kinetic’ seizures report greater pre-seizure sensory symptoms^63^. Conversely the same group have reported that those individuals with the lowest trait dissociation scores have primarily ‘non-hyperkinetic’ seizures^64^. Finally, a separate study has shown that individuals with migraines before or during their FS are more likely to have seizures associated with less movement^65^. The proposed interpretation of our data would also explain why, via shifts in attentional direction, some individuals may have motile and non-motile features within one event. Our findings suggest that seizure semiology may not be random but reflect dynamic interoceptive states. Monitoring HEP or related interoceptive biomarkers could eventually help anticipate seizure expression, inform precision therapeutic targeting, or help differentiate FS subtypes in ambiguous cases.

### Limitations

One limitation of this data is that it lacks contemporaneous behavioural data for many of the patients. While some behavioural information is implied in the annotated symptom reports in the functional syncope recordings, future work should document trait and state interoceptive and autonomic symptomaticity and compare this to the change in HEP. A second limitation the relatively low temporal resolution of the HEP, as it relies on so many averaged heartbeats. This may therefore limit the sensitivity to rapid changes in interoceptive changes. Other methods to index interoceptive processing implicitly, along with behavioural data would further inform the utility of the model. Finally, HRV is an imperfect measure of autonomic activity. Simultaneous beat-by-beat blood pressure allows the computation of carotid sinus sensitivity, which better indexes the sympathetic nervous system, but this again has limited temporal resolution, as well as practical limitations in the context of clinical scenarios. Finally, the findings in this study need to be replicated in a larger sample.

## Conclusion

We have demonstrated that motile and non-motile FS correspond to differences in the change in amplitude of the HEP prior to the seizure. We have used a predictive processing framework to interpret these findings as changes in the precision of the cardiac interoceptive prediction error signalling before a functional seizure. We hypothesise that increased and decreased bodily attention before a non-motile and motile functional seizure respectively lead to the observed semiological differences.

## Data Availability

All data produced in the present study are available upon reasonable request to the authors

## Acknowledgements

The authors would like to thank the staff in the Sir William Gowers Centre for their work in acquiring the clinical data used in this study.

## Data Availability

Data can be made available on request

